# A simple Stochastic model for the SARS-CoV-2 Epidemic curve

**DOI:** 10.1101/2020.05.29.20116723

**Authors:** A. N. Diógenes, D. G. Tedesco

## Abstract

An epidemic curve is a graphic depiction of the number of outbreak cases by date of illness onset, ordinarily constructed after the disease outbreak is over. However, a good estimate of the epidemic curve early in an outbreak would be invaluable to health care officials. On the other hand, from the end of February, the SARS-CoV-2 epidemic in Brazil seems to not follow the Europe, or in particular, Italy or Spain. Even if less tests have been applied, there are less deaths occurring in Brazil than in both cited countries. However, since few tests were applied, there is no certain planning on the real number of active cases. To estimate the number of future cases, epidemiologists make an educated guess as to how many people might become affected. We have proposed a simple fitting model using a simulated annealing technique, testing it with the South Korea data. We have tested and discussed the uncertainties of the model. We also have analyzed the trends in the confirmed cases using this model for the five most affected countries plus Brazil along several epidemic weeks.

## INTRODUCTION

A novel coronavirus disease, namely COVID-19, was first detected in Wuhan city, China in December 2019 Lu et al. (2020) and Xu et al. (2020). In the subsequent months, the virus spread became global Chen et al. (2018), Gilbert et al. (2020) and Sohrabi et al. (2020). To control the spread of COVID-19, several studies have been conducted to explore important factors affecting the transmission of SARS-CoV-2 Wei et al. (2020).

An epidemic is a usual term in epidemiology that refers to the appearance of new cases of a particular disease in a given human population, during a given time period, at a rate that substantially exceeds the expected number based on recent experience Le Strat; Carrat (1999). An epidemic may affect a region, a country, or even a group of countries. If an entire continent or the entire globe is affected, we ordinarily call the occurrence a pandemic, just as the SARS-CoV-2 pandemic situation.

An epidemic curve is a graphic depiction of the number of outbreak cases by date of illness onset. Usually, the time interval is a week, but the data is daily. A good estimate of the epidemic curve during an outbreak would be valuable to health care officials and according to these they can plan for sufficient resources and supplies to handle disease treatment on a timely basis. When we say we are estimating the epidemic curve during an outbreak, we mean that, on a given day of the outbreak, we are estimating the daily number of new cases for days that have occurred so far and we are predicting those daily values for days that will occur in the future. We call the collection of these estimates and predictions the estimate of the epidemic curve. Estimation of an epidemic curve in real-time is quite complex because we need a model of the outbreak (an epidemic model), a model of sickness behavior of individuals, and a model of the surveillance system (any sampling inefficiency, time delays). Several SIR models try to model epidemic situations, having many parameters and uncertainties Huang (2016).

At present, methods for doing real-time estimation and prediction of the magnitude of an outbreak can be very complex, but somehow imprecise. For the most part, investigators simply do their best to intensify surveillance in an effort to identify all cases so that the observed number of cases is as close to the real number of cases as possible Wagner et al. (2006). Several other methods can provide estimates of some outbreak characteristics such as outbreak type, source, and/or route of transmission of the outbreak Ma (2020), Scarabel et al. (2020) and IleVillela (2020). However, none of these estimates the epidemic curve. On the other hand, models like the proposed by Tang Tang et al. (2020), model the epidemic curve, but need several reliable data. Arino (2020) remarks that there is a lot of available data, but there are countries where that is not necessarily true. Knowledge of the probable values of these variables should be more useful to public health officials than merely knowing that an outbreak is probable. However, an estimate of the epidemic curve itself would be better.

The current paper addresses these shortcomings. First, the log-logistic simulated annealing model discussed in this paper estimates the epidemic curve itself. Second, we discuss the uncertainties and challenges when applying the model for Brazilian cities epidemic curves.

## THEORY

The epidemic curve for an outbreak is often correlated with the daily counts of some observable event. A typical SARS-CoV-2 propagation curve can be observed in Figure 1 World meters info (2020a). The South Korea was chosen because the epidemy is said to be controlled, by April 2020. The typical “S” shaped curve indicates that the propagation follows both an exponential trend, but sometime it changes to an almost linear trend. Therefore, regular exponential fits do not work, and the challenge is to predict the exponential decay during the curve (Ma, 2020).

**Figure 1.**
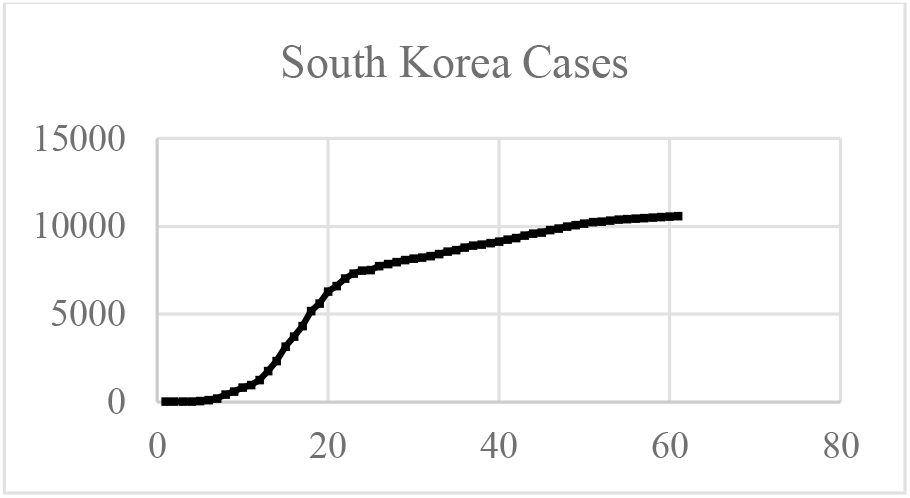
South Korea SARS-CoV-2 total cases

The proposed model uses a flexible analogical equation to model the epidemic curve and predict the entire curve for a time period.

### Simulated Annealing

Simulated Annealing is the probabilistic meta-heuristic adopted in this work and it was chosen due to its capacity of “escape” from local minima (which are very frequent on this problem). It is also worth of mention that the process of recrystallisation, the inspiration for simulated annealing, is a natural instance of a placement problem (Kirkpatrick et al. 1983).

### Description

Simulated annealing comes from the Metropolis algorithm, a simulation of the recrystallisation of atoms on a metal during its annealing (gradual and controlled cooling). During annealing, atoms migrate naturally to configurations that minimize the system total energy, even if during this migration the system must pass through high-energy configurations (Kirkpatrick et al. 1983).

The observation of this behavior suggests the application of the simulation of such process to combinatorial optimization problems. Simulated annealing is a hill-climbing local optimization heuristic, which means it can skip local minima by allowing the exploration of the space in directions that lead to an increase on the cost function. It sequentially applies random modifications on the evaluation point of the cost function. If a modification yields a point of smaller cost, it is automatically kept. Otherwise, the modification also can be kept with a probability obtained from the Boltzman distribution (Kirkpatrick et al. 1983).

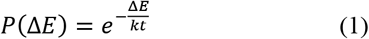

where P(ΔE) is the probability of the optimization process to keep a modification that incurred on an increase ΔE of the cost function, k is a parameter of the process (analogous to the Stefan-Boltzman constant) and t is the instantaneous “temperature” of the process. This temperature is defined by a cooling schedule, and it is the main control parameter of the process. Several cooling schedules can be evaluated to solve a problem, as an adaptive cooling (Martins; Tsuzuki, 2005).

The adopted in this research was the adaptive cooling used by Martins and Tsuzuki (2005). Likewise, the initial temperature was chosen as Heckmann & Lengauer (1995) proposed.

### Log-Logistic curve

The log-logistic distribution is a continuous probability distribution whose logarithm has the logistic distribution pioneered to model population growth (Verhulst, 1838). This model is well-known as the Fisk distribution and is often used to model random lifetimes in many areas such as reliability, survival analysis, actuarial science, economics, engineering. In some cases, the log-logistic distribution is a good alternative to the log-normal distribution, since it characterizes increasing hazard rate function (HRF). Its equation is very versatile and specifically its cumulative distribution function can have a “S” shape, as exposed in Figure 2.

**Figure 2.**
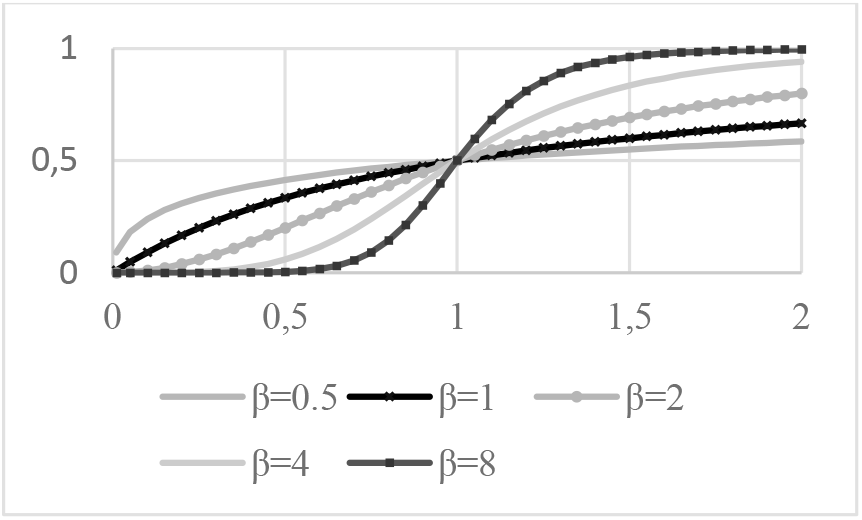
Cumulative distribution function for the log-logistic curve for α = 1 and varying β.

The equation for this curve is exposed in (2).

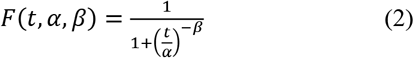

Other curves have similar shapes, like the Generalized extreme value distribution, or Fisher-Tippet distribution (20). However, Diógenes et al. (2011) already used the Log-Logistic distribution successfully to model grain size distribution curves.

## RESULTS AND DISCUSSION

The Brazilian Institute for Geography and Statistics (IBGE) makes available the COVID-CoV-2 propagation data for every city that has cases in Brazil. The selected cities were the 5 most affected until April 16th plus Brasília, Brazil’s capital. The data was collected and processed using a simulated annealing technique to fit a log-logistic curve.

Since the log-logistic distribution curve has its maximum in the 1 value, it was added a multiplier that represents the limit of infected persons along time. Therefore, the simulated annealing method is varying three parameters: both the α and β from the regular log-logistic distribution, but also the multiplier which will be the infected person limit.

### Model reproducibility

The South Korea curve was run five times to evaluate the model reproducibility. South Korea data was chosen because this Country tested massively its population and its data is said to be reliable. The results for the five simulations can be observed in Figure 3.

**Figure 3.**
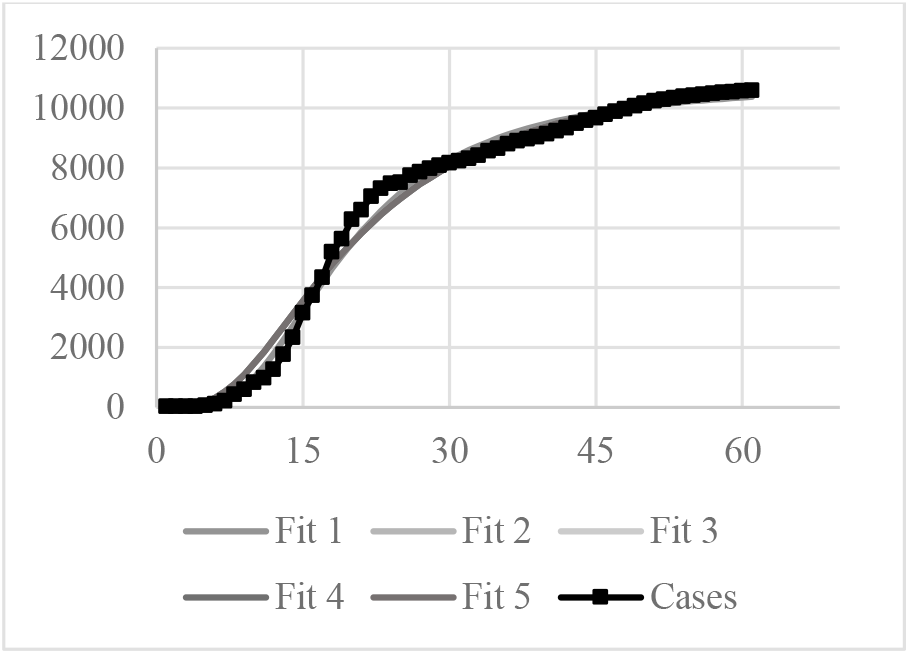
Five Log-logistics fit compared to South Korea confirmed cases

The estimated maximum infected population was 10,909 for the Fit 1 and 11,650 for the other four simulations.

### Brazilian cities data fitting

The Brazilian cities selected were five more affected plus the capital. The Brazilian Institute of Geography and Statistics – IBGE (2020) provides several data about the selected cities. These data are exposed in Table 1. The number of tests per million of habitants for Brazil is 3462 (World meters info 2020b).

**Table 1.** Data for selected cities

| City | Population (Millions of hab.) | Demographic density (hab./km<sup>2</sup>) |
| --- | --- | --- |
| São Paulo | 11.253 | 7,3328.26 |
| Rio de Janeiro | 6.320 | 5,265.82 |
| Fortaleza | 2.452 | 7,786.44 |
| Manaus | 1.802 | 158.06 |
| Recife | 1.537 | 7,039.64 |
| Brasília | 2.570 | 444.66 |

A typical result is exposed in Figure 4. This figure shows both the fit, but also the plotted fit from the first confirmed case date to 04/30. The black line represents the confirmed cases, while the grey line represents the simulated annealing log-logistic model. The R^2^ for each fit is exposed in Table 2.

**Table 2.**
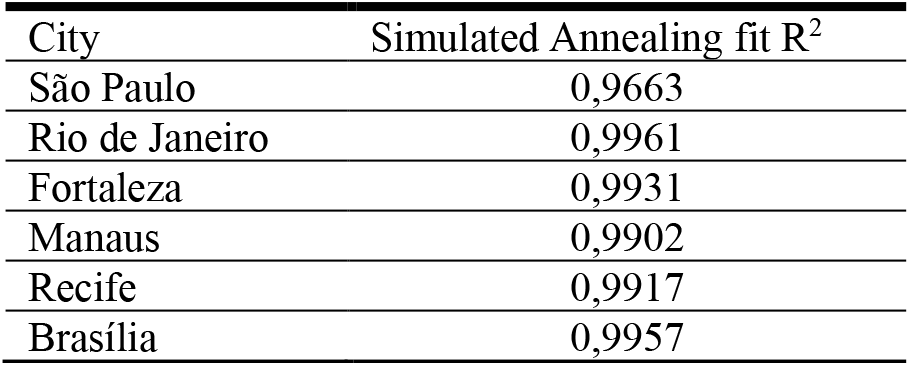
R^2^ for the simulated annealing log-logistic fit City.

| City | Simulated Annealing fit $R^2$ |
| --- | --- |
| São Paulo | 0,9663 |
| Rio de Janeiro | 0,9961 |
| Fortaleza | 0,9931 |
| Manaus | 0,9902 |
| Recife | 0,9917 |
| Brasília | 0,9957 |

**Figure 4.**
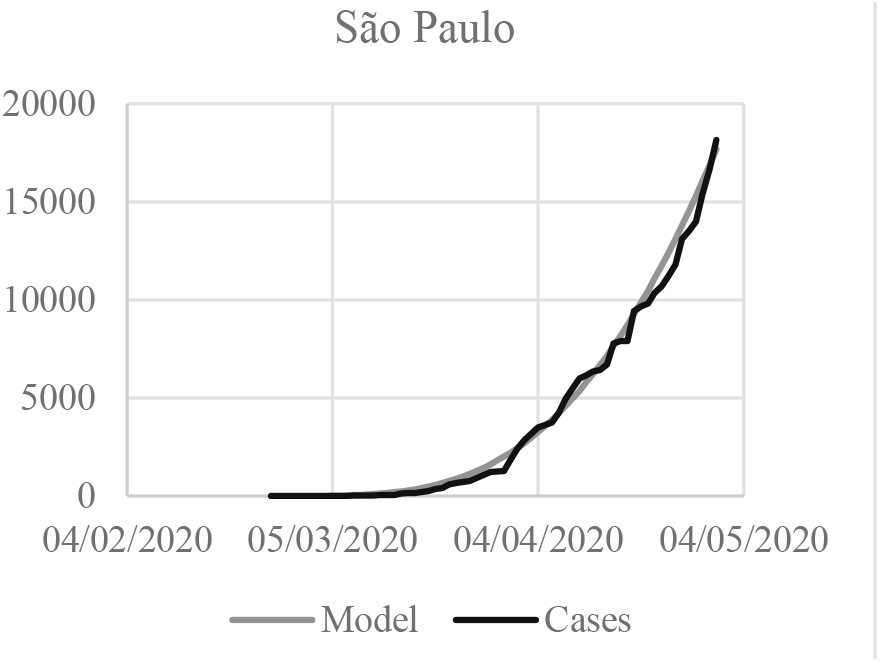
(a) Log-Logistic fit for São Paulo with the data from 02/25 to 04/30.

The other simulated cities showed similar results. There are two important information that can be obtained from the model: the maximum rate of contagious and the number of infected people at 12/31/2020. This information is exposed in Table 3 for the analyzed cities.

**Table 3.**
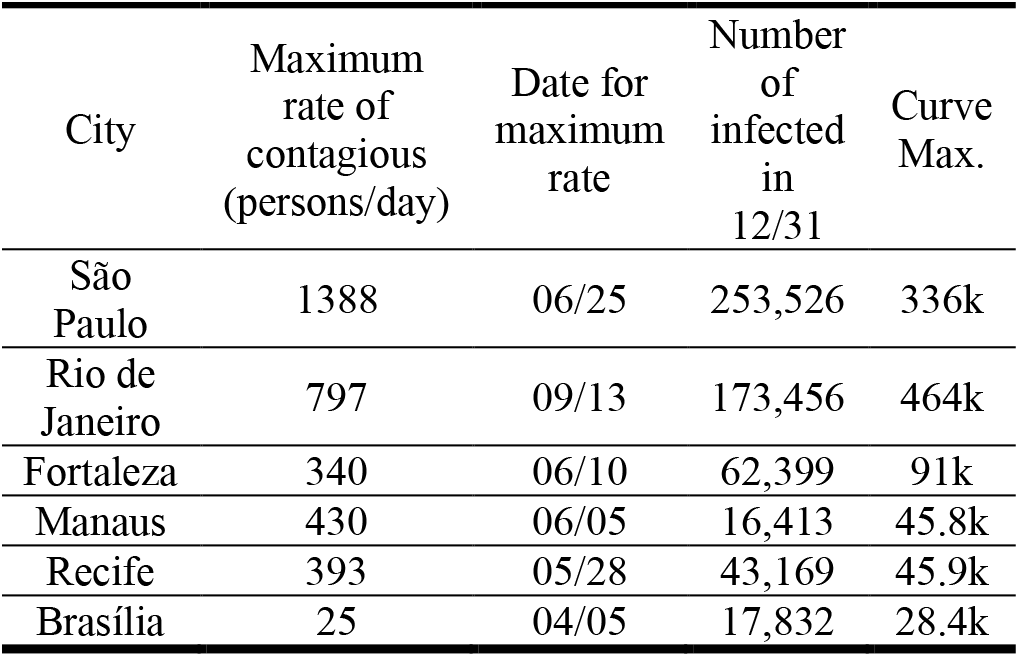
Maximum rate and number of infected people for the analyzed cities

| City | Maximum rate of contagious (persons/day) | Date for maximum rate | Number of infected in 12/31 | Curve Max. |
| --- | --- | --- | --- | --- |
| São Paulo | 1388 | 06/25 | 253,526 | 336k |
| Rio de Janeiro | 797 | 09/13 | 173,456 | 464k |
| Fortaleza | 340 | 06/10 | 62,399 | 91k |
| Manaus | 430 | 06/05 | 16,413 | 45.8k |
| Recife | 393 | 05/28 | 43,169 | 45.9k |
| Brasília | 25 | 04/05 | 17,832 | 28.4k |

### Brazil data fitting

A Brazilian as country fitting was also performed. The resulting fit is exposed in Figure 5.

**Figure 5.**
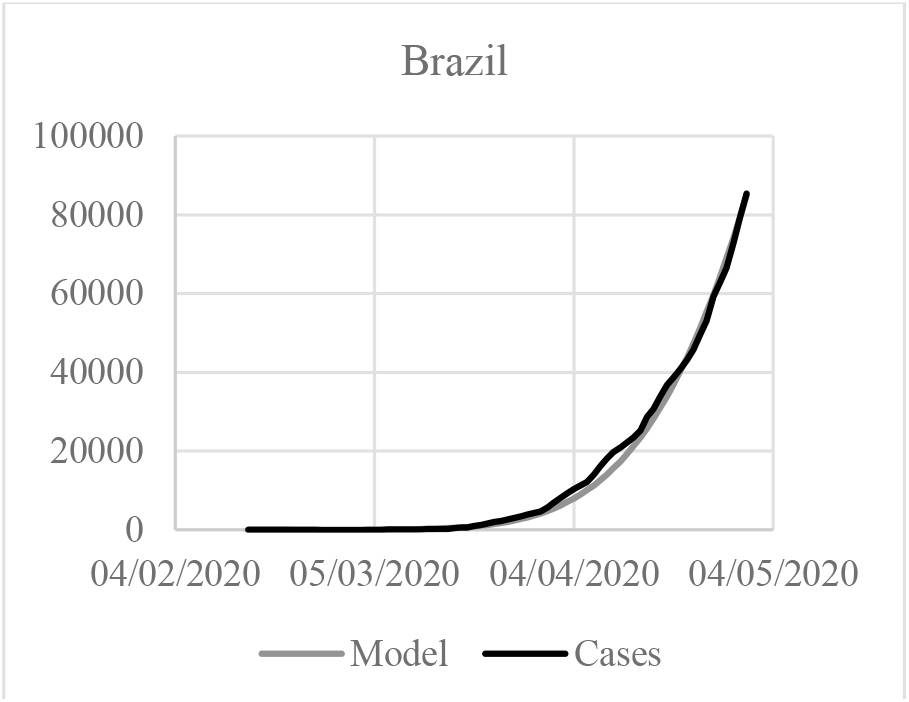
(a) Log-Logistic fit for Brazil with the data from 03/07 to 04/30.

The log-logistic fit models provided interesting information. All curves were properly fit, however all of them with different behaviors, which are compatible with the cities’ realities.

São Paulo is Brazilian greatest city, and also one of the most populated in the world. Likewise, it was the first Brazilian city to have SARS-CoV-2 cases. Therefore, the curve “S” shape is more visible and also easier to fit. The curve trend goes to 336k of infected persons, but the 2020 terminates with only 253.5k cases.

The quadratic error was the lesser of the analyzed cities. That was caused by fluctuations in the curve. These fluctuations come from the new cases curve. At São Paulo, it has big variations, as it can be observed in Figure 6. The confirmed cases (blue varying line) curve varies a lot. That happens due a lot of factors, but one significant factor is that the Brazilian laboratories that process the tests do not have a constant work rate, therefore some days have more published cases than others, but that does not mean that the contagion rate has changed. It just means that a confirmed case was not published in a day, rather it was published in the following days, however that affects the curve fitting quality.

**Figure 6.**
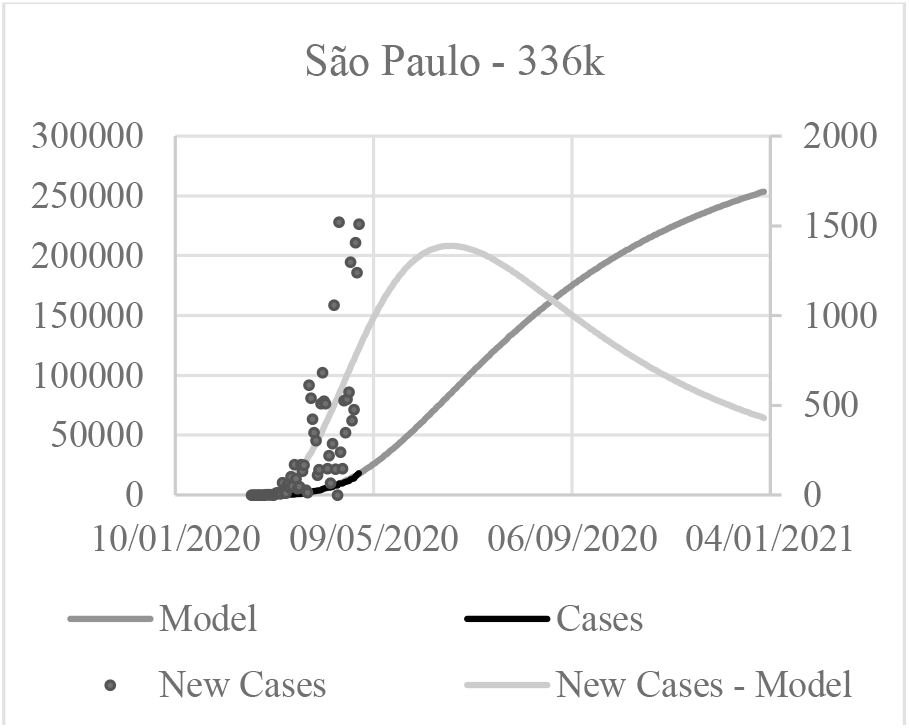
Comparison of the New cases curve for the model and the confirmed cases for São Paulo

Rio de Janeiro had a particular good fitting, and the estimated infected peak population was greater than the São Paulo. That can be merely a statistical error, since the Rio de Janeiro population is smaller. Otherwise, the Rio de Janeiro population has many favelas, which makes the population very concentrated and also susceptible to the SARS-CoV-2 propagation. However, that factors were not considered in the analysis. The propagation after May seems to be almost linear.

Fortaleza and Manaus data had similar fits and both of the cities tend to have the peak propagation for June. Recife seems to be slightly different, as the curve fitting seems to indicate a faster propagation, but also a faster peak. Also, Recife seems to change the curve fitting by April 20th. The explanation for this behavior is that from April 22nd to April 24th there was no new cases, which changed the curve trend. The authors choose to keep the fit model.

Brasília was an interesting case. Even with an excellent fitting, since the SARS-CoV-2 propagation until April 12th was almost linear, the infected population for the curve fitting has a different behavior. A possible cause for this is the massive fluctuation on the new confirmed cases real data. The fluctuations are so scattered, that it was not possible to accurately model it, and also the maximum rate of contagious was smaller in the model than the real data. Another possible explanation is that Brasília population is the most spread across the analyzed cities, however that issue was not considered in the fit. The Brazil curve fitting had a similar behavior to São Paulo. That is adequate, because both city and country have the most significant data. Also, the Brazil fitting maximum population is consistent with the other fittings.

The fitting model has its limitations, since it does not consider population parameters, as age, geographical distribution, death rate, morbidities or any other parameter than the curve itself. However, even with these limitations, the model can be a valuable tool, since it helps to understand in a very simple way how the SARS-CoV-2 is spreading using simple computational tools and almost no data at all. Likewise, with simple data interpretation, the model can be very useful.

Since in Brazil the test quantities are very limited, it is possible to use this methodology to configure a transfer function to estimate the real epidemic curve based on the death cases. That issue shall be studied in a near future

## CONCLUSIONS

Our study suggests that a model with simple computational tools and with strong limitations in the feeding data can be useful to model the SASR-CoV-2 virus spreading across a population. The model was performance was evaluated with South Korean data and it was applied to six Brazilian cities. The model helped to understand several uncertainties and challenges on the SASR-CoV-2 epidemic curve model. Further studies are required to understand the model potential.

## Data Availability

The authors confirm that the data supporting the findings of this study are available within the article and its supplementary materials.

## ACKNOWLEDGEMENTS

The authors acknowledge with gratitude the support of the Universidade Positivo.

## NOMENCLATURE

E: energy
k: Bolzmann constant
P: probability
R: correlation parameter
t: time
T: temperature

## Greek symbols

α: distribution parameter
β: distribution parameter
Δ: variation

